# Severe preeclampsia is not associated with significant DNA methylation changes but cell proportion changes in the cord blood - caution on the importance of confounding adjustment

**DOI:** 10.1101/2023.08.31.23294898

**Authors:** Wenting Liu, Xiaotong Yang, Zhixin Mao, Yuheng Du, Cameron Lassiter, Fadhl M. AlAkwaa, Paula A Benny, Lana X Garmire

**Author notes:** These authors contributed equally to the work (Wenting Liu, Xiaotong Yang).

## Abstract

Epigenome-wide DNA methylation analysis (EWAS) is an important approach to identify biomarkers for early disease detection and prognosis prediction, yet its results could be confounded by other factors such as cell-type heterogeneity and patient characteristics. In this study, we address the importance of confounding adjustment by examining DNA methylation patterns in cord blood exposed to severe preeclampsia (PE), a prevalent and potentially fatal pregnancy complication. Without such adjustment, a misleading global hypomethylation pattern is obtained. However, after adjusting cell type proportions and patient clinical characteristics, most of the so-called significant CpG methylation changes associated with severe PE disappear. Rather, the major effect of PE on cord blood is through the proportion changes in different cell types. These results are validated using a previously published cord blood DNA methylation dataset, where global hypomethylation pattern was also wrongfully obtained without confounding adjustment. Additionally, several cell types significantly change as gestation progress (eg. granulocyte, nRBC, CD4T, and B cells), further confirming the importance of cell type adjustment in EWAS study of cord blood tissues. Our study urges the community to perform confounding adjustments in EWAS studies, based on cell type heterogeneity and other patient characteristics.

## Introduction

DNA methylation is a type of epigenetic modification that plays a crucial role in regulating gene expression and maintaining genome stability. It involves the addition of a methyl group to the cytosine base of DNA, typically at the CpG dinucleotide sites^1^. Methylation at these sites can affect gene expression by altering the accessibility of DNA to transcription factors and other proteins. DNA methylation profile can be modified by various factors, including aging, diseases, and environmental changes^2,3^. Previous studies on DNA methylation have contributed to biomarker identifications for risk prediction, early detection, and prognosis tracking of various diseases^4–6^. Differential methylation analysis, or epigenome-wide association (EWAS) study, is a key computational process to identify disease-associated DNA methylation markers^7^. However, rigorous statistical and bioinformatics approaches remain a central issue to draw unbiased conclusions in these studies. Recently, some researchers have started to raise the awareness of confounding adjustment for DNA methylation results, including clinical characteristics and heterogeneity of cell types within sample tissues^8^. Failure to properly adjust for these confounders may lead to biased and inaccurate results.

In this study, we alert the community to the importance of confounder adjustment, using the case study of the DNA methylation change in cord blood samples from babies born of severe preeclampsia (PE). PE is one of the leading causes of maternal and prenatal morbidities and mortalities, affecting 2-8% of pregnancies globally and around 3.1% in the US^9,10^. PE is characterized by new-onset hypertension with proteinuria or one/more adverse conditions after 20 weeks of gestation^11^. It can lead to severe outcomes including renal failure, seizure, multiorgan dysfunction, and stroke in mothers; as well as intrauterine growth restriction (IUGR) and premature delivery of the fetuses. Based on blood pressure, clinical findings, and degree of proteinuria, PE can also be classified into severe PE or mild PE. Severe PE poses a greater risk to maternal and fetal health and may involve different pathways than mild PE of similar onset time^12^. It is imperative to investigate the molecular mechanisms of severe preeclampsia.

Many previous epigenetic-wide studies aim to find biomarkers of PE using complex tissues, such as placenta tissues^13,14^, maternal gestational blood^15^, and fetal chorioamniotic membrane^16^. Multiple previous studies looked into the effect of PE on cord blood^17–20^. Yet most of these earlier studies didn’t adjust for potential confounding factors using either cell-type heterogeneity^17–19^ or clinical variables, such as gestational age another significant confounder for pregnancy diseases including PE^20^. Tissues like placentas and cord blood consist of many diverse cell types, each with a distinct epigenetic profile as defined^21,22^. Thus the varying cell types in each sample can affect the overall DNA methylation profile at the bulk level. It is therefore essential to account for such heterogeneity, to improve the accuracy and sensitivity and avoid wrongful conclusions of EWAS biomarker detection. Particularly, to ensure that any differences in DNA methylation are due to confounding factors, the analysis needs to be adjusted for cell proportions. Moreover, if the essential clinical data such as gestational ages are available (as they should be), then the analysis needs to be adjusted for the important clinical variables as well. We show the practice of doing such cell type and clinical confounding adjustment using the case study of cord blood DNA methylation analysis. We warn the community of potential significant harm otherwise.

## Materials and Methods

### Study cohort

The umbilical cord whole blood DNA samples were obtained from Hawaii Biorepository (HiBR). The HiBR collected placenta, maternal, and cord blood samples from deliveries at Kapiolani Women and Children’s Hospital from 2006 to 2013. It is one of the largest research tissue repositories in the Pacific region, containing specimens from more than 9250 mother-child pairs at the time of sample collection. Umbilical cord samples were collected immediately after delivery. Severe PE was characterized by OBGYNs at Kapiolani Medical Center as sustained pregnancy-induced hypertension(systolic/diastolic blood pressure >= 140/90) with urine protein and/or organ dysfunction. We originally collected 63 samples. The demographic and clinical information of the patients was collected and analyzed to identify any potential confounding effects. Data usage was approved by IRB #CHS23976.

### Sample preparation

Umbilical cord blood samples were collected in the operating room immediately after delivery. To prepare for DNA extraction, we first added three volumes of RBC Lysis Solution to one volume of clotted blood, which is then vortexed and incubated on a shaker for 15 minutes at room temperature. The sample was then centrifuged to pellet white blood cells and clot particulates, and the supernatant is carefully poured into a waste bucket. The pellet was resuspended in an additional volume of RBC Lysis Solution and incubated again for 15 minutes. After another centrifugation step, the supernatant was carefully removed, leaving behind 200 µL of residual liquid. The pellet was then vigorously resuspended in the residual liquid before being combined with a master mix of Cell Lysis and Proteinase K Solution. The mixture was vortexed and incubated at 55°C until homogenous, with intermittent vortexing to facilitate digestion. Once homogenous, the samples were subjected to DNA purification on the Autopure Machine following the manufacturer’s instructions.

### DNA extraction and methylation profiling

DNA was extracted from prepared cord blood samples using AllPrep DNA/RNA/Protein Mini Kit (Qiagen, USA) according to the manufacturer’s instructions by HiBR. We obtained pre-extracted genomic DNA of whole cord blood samples from the HiBR and conducted DNA Illumina EPIC Beadchip assays through the University of Hawaii Cancer Center Genomics Core.

### DNA methylation data pre-processing and quality control

We used the R package “ChAMP” for data pre-processing (**Supple Fig. 1).** We first filtered probes using the following criteria sequentially: (1) removing probes with a detection p-value above 0.01 (7,941 probes); (2) removing probes with a bead count <3 in at least 5% of samples (27,731 probes); (3) removing probes with no matched CpG sites (2,673 probes); (4) removing probes that align to multiple locations (8,248 probes). During the quality control step, we removed 1 control sample with a distinct beta density distribution (**Supple Fig. 2A, 2B**). We normalized the remaining samples using BMIQ methods^23^, and corrected for batch effects using the ComBat algorithm embedded in “ChAMP”. We used the singular vector decomposition (SVD) heatmap to verify the effectiveness of batch removal (**Supple Fig. 2C, 2D**). The preprocessed data matrix contains 62 samples and 819,325 probes. We converted the original methylation intensity (beta) to M-values using “beta2m” function from “lumi” package to reduce heteroskedasticity^24^, where M-values are defined as the log2 ratio of the beta value of each probe.

**Fig. 1:**
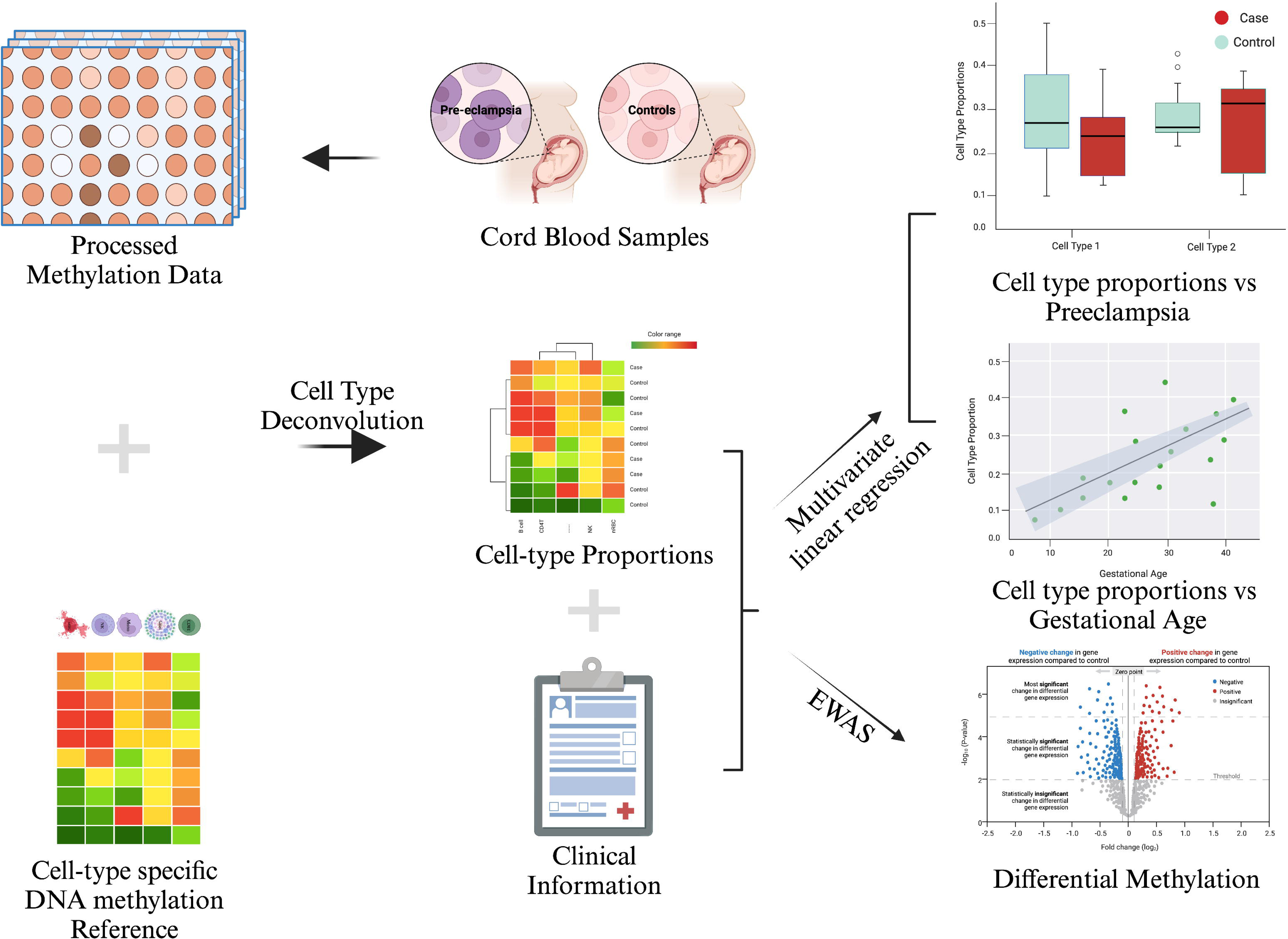
Study Overview and experiment design. The entire data analysis procedure is outlined in this workflow, which incorporates methods that account for clinical confounding and cell type confounding.

**Fig. 2:**
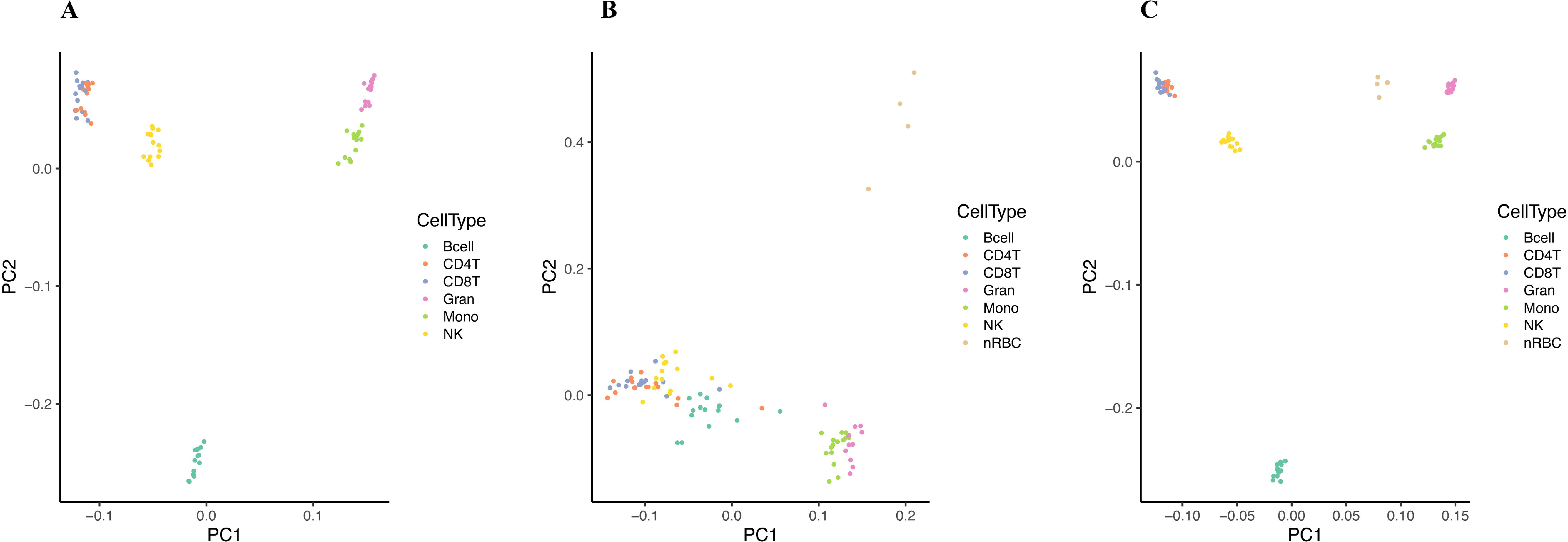
Construction of cell type reference matrix. Principal Component Analysis (PCA) plots of DNA methylation reference matrix from (A) Lin et al.^25^, (B)Bakulski et al.^26^, and (C) our merged reference, colored by cell type. Our reference combined nucleated red blood cells (nRBCs) from Bakulski et al and 6 cell types (B cell, CD4T, CD8T, monocyte, natural killer cell, and granulocyte) from Lin et al^25^. Our reference shows better separation between cell types, compared to the references of Lin et al^25^. and Bakulski et al.^26^

### Cell-type deconvolution in umbilical cord whole blood (CB)

Bulk-level DNA in umbilical cord whole blood (CB) includes at least 7 most common blood cell types: neutrophils, B cells, CD4T, CD8T, monocytes, natural killer cells (NK), and nucleated red blood cells (nRBC). Each sample may have different compositions of the cell types above, thus needing deconvolution. For this, we combined two methylation reference matrices of the cord blood sample for better cell-proportion estimation from the whole blood samples. We extracted a reference panel of B cell, CD4T, CD8T, monocyte, natural killer cell, and granulocyte from Lin et al.^25^ given the reported high quality of the reference samples. We also added the DNA methylation profiles of nucleated red blood cells (nRBC) from Bakulski et al^26^ to be part of the whole blood DNA methylation reference. We used the “combineArrays” function in the “minfi” R package to combine two data sets and rescaled them using the “BMIQ” method in the “wateRmelon” R package. We used pairwise t-tests with Bonferroni adjustment (threshold of 1E-8) to identify significant CpGs among the 7 cell types in each pair as cell-specific markers, similar to others^25^. This process yielded 151,794 cell-type specific CpG biomarkers for the DNA methylation reference matrix for deconvolution. We uploaded the new whole blood CpG reference dataset in the EpiDISH package.

We used principal component analysis (PCA) to check the quality of these new sets of markers. We applied the selected markers above to estimate the cell type proportions in each sample, using the reference-based cell-type deconvolution algorithm Constrained Projection (CP) from the “EpiDISH” package^27^. We used the resulting cell type proportions to adjust for confounding effects in the epigenome-wide association analysis.

### Clinical confounders and source of variance analysis

We retrieved a total of 6 commonly reported clinical variables in cord blood EWAS studies from the biobank, including maternal age, ethnicity, parity, BMI, delivery gestational age, and smoking status^20^. We imputed 3 samples (including 1 severe preeclampsia and 2 controls) with missing BMI using mean values of each sample group. We performed the source of variance (SOV) analysis on the 6 clinical variables and previously estimated sample cell proportion to identify important confounding variables that need to be adjusted, as done before^28,29^. SOV analysis calculates F-statistics that can be used to identify and quantify the contribution of different factors to the total variance in a dataset. We adjusted the variables with F-statistics larger than 1 (the error value) as confounding variables.

### CpG-level epigenetic-wide association analysis (EWAS)

We calculated the differentially methylated probes (DMP) between severe PE cases and controls using moderated t-test with Benjamini-Hochberg (BH) adjustment (threshold of 0.05). We included study participants’ gestational age (GA), BMI, parity status, ethnicity, and methylation-derived cell compositions in the linear model to remove the confounding effects, and compared the result with that without confounding adjustment using “limma” package^30^. We defined hypermethylated CpGs as significant CpGs with positive log2-transformed fold change (logFC) and hypomethylated CpGs as significant CpGs with negative logFC, respectively. We used volcano plots to illustrate the global DNA methylation changes between the cases and controls.

### Gene-level EWAS

We further examined the methylation signal difference between severe PE and controls at gene and pathway levels. We annotated the CpGs^31^, selected those located on the promoter region and aggregated the methylation signals of all CpGs within a gene promoter by taking the geometric mean. Then we compared the aggregated methylation signals between severe PE cases and control groups, using the moderated t-tests with BH adjustment. Additionally, we looked for pathways associated with promotor region methylation differences using the R package pathifier^32^. The pathifier algorithm calculates a pathway deregulation score (PDS) for each sample and each pathway. We compared the pathway PDS scores in case and control samples by moderated t-tests with Benjamini-Hochberg FDR adjustment (threshold p-values 0.05).

### Software Usage and Code Availability

All analysis was done using R 4.1.2^33^. Specially, we used “ChAMP” (version 2.24.0) for data preparation^34^, “limma” for differential analysis^30^, “EpiDISH” (version 2.10.0) for cell-type deconvolution^27^, and “IlluminaHumanMethylationEPICanno.ilm10b4.hg19” for data annotation^31^. All codes are available at https://github.com/lanagarmire/CB_DNAm_PE.

## Results

### Overview of Study Design and Cohort Characteristics

The overview of the study design is illustrated in **Fig. 1**. We obtained whole cord blood samples from 24 severe PE cases and 39 controls collected at Hawaii Biorepository (HiBR) from January 2006 and June 2013. The maternal demographic and clinical characteristics are shown in **Table 1**. There is no significant difference (P > 0.05) in maternal age, parity, BMI, ethnicity, and smoking status. Noticeably, the severe PE cases have earlier delivery gestational age compared to healthy controls (P = 3.66E-06), as the clinical management of severe PE usually often demands early delivery to avoid severe maternal morbidities. We obtained pre-extracted genomic DNA of whole cord blood samples from the HiBR and conducted DNA Illumina EPIC Beadchip assays through the University of Hawaii Cancer Center Genomics Core. We conducted bioinformatics pre-processing of the DNA methylation following standard steps in R package “ChAMP” (**Supple Fig. 1**). Briefly, we first filtered out probes of bad quality, then examined the methylation level distribution of each sample and removed 1 control sample with abnormal distribution (**Supple Fig. 2A, 2B**). Lastly, we normalized the data and removed the batch effect between arrays (**Supple Fig. 2C, 2D**). The remaining preprocessed data matrix contains 62 samples and 819,325 probes (see **Methods**).

**Table 1:** Patient Characteristics.

| *Variables* | *PE Cases*<br>*(n = 24)*<br>*mean (sd)* | *Controls*<br>*(n = 38)*<br>*mean (sd)* | *P-value* |
| --- | --- | --- | --- |
| *Maternal Age (Years)* | 28.75 (5.88) | 27.24 (6.35) | 0.34 |
| *Parity* | 1.54 (1.41) | 1.57 (1.78) | 0.93 |
| *BMI* | 32.24 (9.38) | 27.75 (9.20) | 0.07 |
| *Smoker (n)* | 6 | 8 | 0.96 |
| *Gestational Age (Weeks)* | 35.58 (2.90) | 39.16 (0.92) | 3.66E-06 |
| *Ethnicity (n)* |  |  | 0.60 |
| *Asian* | 12 | 21 | - |
| *Caucasian* | 3 | 7 | - |
| *Pacific Islander* | 9 | 10 | - |

\* Numeric variables are compared with *t*-test;
\* Categorical variables are compared with the Chi-square test.

### Cell-type deconvolution in cord blood samples

Cord blood is a complex tissue with various cell types, including granulocyte, B cell, CD4T, CD8T, monocyte, natural killer, and nucleated red blood cells (nRBC), each with a unique DNA methylation profile. Cell type heterogeneity in the cord blood could significantly confound the phenotypes of interest, affecting the differential methylation analysis results. Previous studies showed that cell type heterogeneity contributes to much more variation in DNA methylation in various tissues, compared to ethnicity, sex, age, and even phenotypes of interest^8,35,36^. If left unadjusted, such variation from cell type difference will result in biased and even misleading results in the differential methylation analysis associated with the phenotype. Therefore adjusting for cell type heterogeneity is an essential step in epigenetic-wide association studies (EWAS) of the bulk-level data.

To adjust for cell-type heterogeneity, we first estimated the cell-type proportions in the cord blood samples by performing cell-type deconvolution. Unfortunately, not all cord blood cell-type specific references contain nucleated red blood cells (nRBC), whose amount decreases as gestation progresses and can be non-trivial in severe PE where most deliveries are preterm^37–39^. To address this limitation, we created a new whole cord blood DNA methylation reference by merging two existing references: the EPIC array reference panel by Lin et al.^25^ and the 450K array reference by Bakulski et al^26^. Lin’s work demonstrated a superior quality of cell type-specific markers, as evidenced by better cluster separation in PCA analysis, compared to Bakulski’s data (**Fig. 2A, 2B**). However, Lin’s reference lacked the crucial nRBC cell type. Therefore, we combined the methylation levels of B cell, CD4T, CD8T, monocyte, natural killer cell, and granulocyte in Lin’s reference and nRBC from Bakulski’s reference. We processed the CpG markers similar to the original studies (see **Methods**). As a result, we identified 151,794 differentially methylated CpGs as high-quality markers for the whole cord blood. The PCA result of the combined reference panel confirmed better separation of cell types than those from both original studies (**Fig. 2C**).

### Association between cord blood cell type and severe PE

We next aimed to determine the significance of cell type heterogeneity as a potential confounder that should be adjusted for in the study. To learn the association between severe PE and cord blood cell composition, we first performed cell-type deconvolution on cord blood samples using the new combined reference and Houseman’s constrained projection(CP) deconvolution algorithm^40^. Estimated cell-type proportions show large variations across samples, especially for granulocyte, nRBC, and CD4T cells (**Fig. 3A**). Granulocyte proportions are significantly lower (□=-0.10, P=3.44e-7) in the severe PE group compared to the control group, while B cell (□=0.0085, P=0.011), nRBC cell (□=0.062, P=1.67e-5) and CD8T cell (□=0.014, P=0.0084) proportions are significantly higher in cases (**Fig. 3B**).

**Fig. 3:**
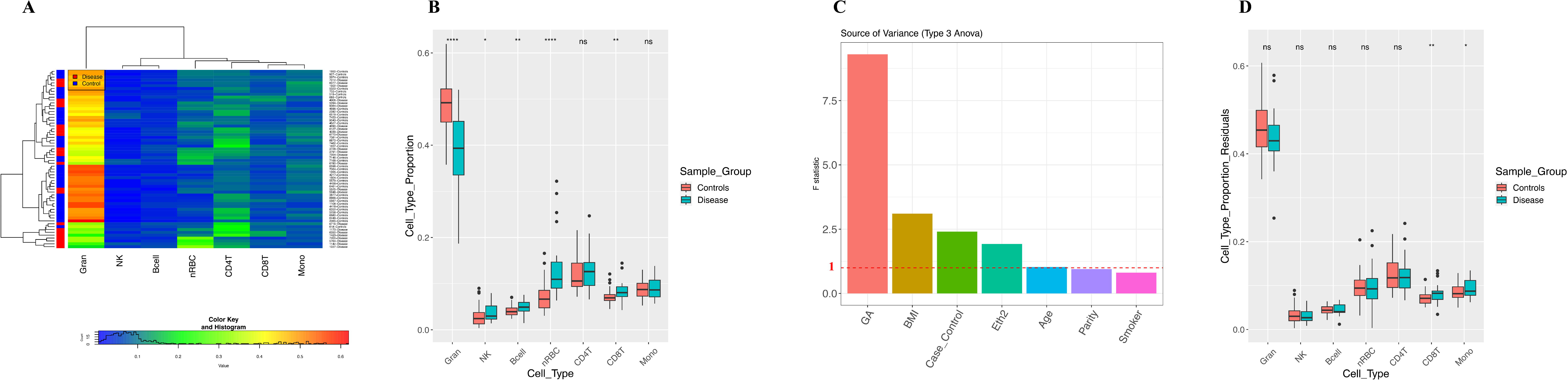
Cell types in samples. (A) Heatmap displaying estimated cell-type proportions among 62 samples (including 24 PE cases and 38 controls), the colors indicate the relative proportions of cell types, with red indicating a higher proportion and blue indicating a lower proportion. (B) Side-by-side boxplots displaying cell-type proportions in PE cases vs. controls before adjusting for clinical variables. An asterisk (*) is used to indicate a significant difference by using Multiple Linear Regression (MLR) between the case and control groups (p-value < 0.05), while "ns" is used to indicate a non-significant difference. (C) The Source of Variance (SOV) analysis of cell type composition from patient characteristics. Confounding factors were identified by considering variables with an F-mean value greater than 1. (D) Side-by-side boxplots displaying cell-type proportions in PE cases vs. controls after adjusting for confounders identified in (C). An asterisk (*) is used to indicate a significant difference by using Multiple Linear Regression (MLR) between the case and control groups (p-value < 0.05), while "ns" indicates a non-significant difference.

However, the apparent differences in cell proportions in cases vs controls could be very well due to other reasons (eg. gestational age) rather than severe PE. To confirm this speculation, we conducted the source of variance (SOV) analysis of cell type compositions on the clinical variables and ranked them by F-statistics. A variable with F-statistics bigger than 1 (the error term) is considered a significant contributor to cell proportion variations. As shown in **Fig. 3C**, in addition to severe PE, gestational age, maternal BMI, and ethnicity also contribute significantly to cell type heterogeneity. Gestational age and maternal BMI rank higher than severe PE. With such caution, we adjusted confounding by linearly regressing cell type proportions over severe PE and other covariate factors, including gestational age, maternal age, ethnicity, BMI, and smoking status. We plot the cell proportions in severe PE vs. controls, post-adjustment by other clinical variables (**Fig. 3D**). The previously observed differences in granulocytes, B cells, and nRBCs now all disappear. CD8T proportions, however, continue to be significantly higher in severe PE cases (□=0.02, P=0.0068). Interestingly, monocyte proportions, which were not significantly different before adjustment between cases and controls (**Fig. 3B**), now become significantly associated with severe PE (□=0.016, p=0.028) after confounding adjustment. The detailed linear regression results of cell proportion on clinical data can be found in **Supple Table 1.** In all, these results show that cell type proportions vary among newborns and it is important to adjust for potential confounding before interrogating the association with severe PE.

### Confounding adjustment drastically affects EWAS results

Considering the current state of most EWAS studies which often overlook the adjustment of cell types and other clinical covariates (such as gestational age) within their samples, we further investigated the impact of these factors on the differential methylation analysis. We conducted the source of variance (SOV) analysis of the DNA methylation matrix on cell type proportion and clinical variables. Strikingly, all cell-type composition variables show the strongest and most dominant explanation power of variation in the methylation data (**Fig. 4A**), ranking even higher than the severe PE condition. After the cell proportions, severe PE case/control, gestational age, maternal age, parity, and ethnicity also have larger F-statistics than the error term (F-statistics=1), in descending order. Therefore, in the downstream analysis of differentially methylated CpGs, we adjust these variables for confounding effects.

**Fig. 4:**
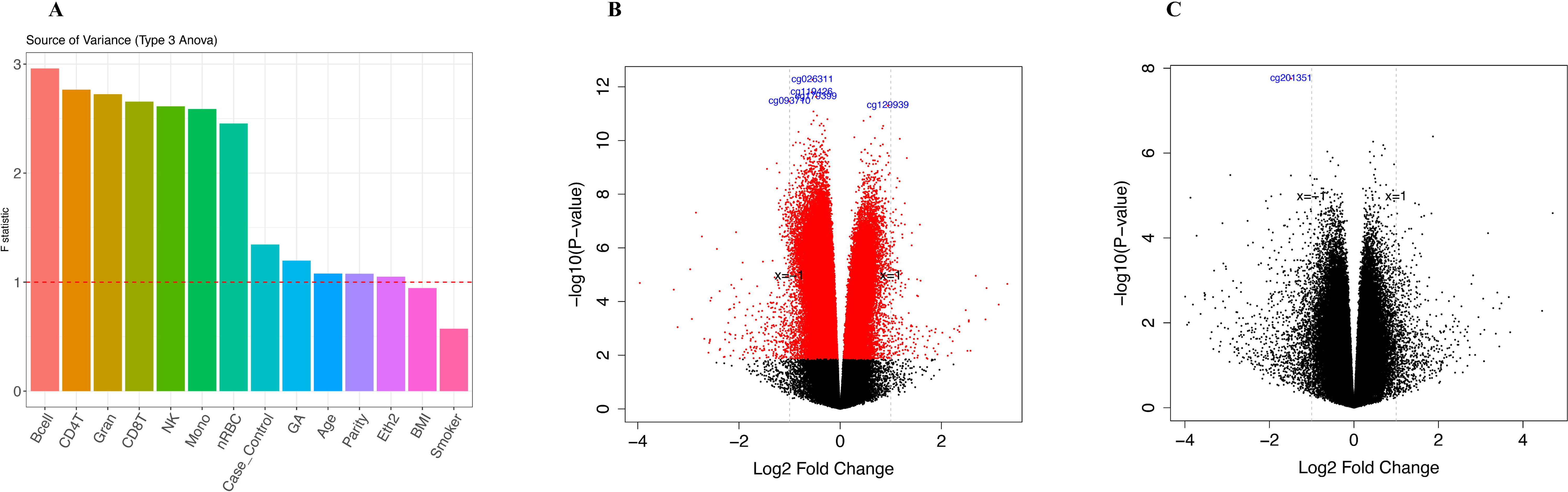
PE is not associated with significant changes in cord blood DNA methylation, after confounding adjustment. (A) The Source of Variance (SOV) analysis was conducted on both clinical variables and cell types. Confounding factors were identified by considering variables with an F-mean value greater than 1. (B) The volcano plot of the differential methylation analysis results without confounding adjustment. The x-axis represents log fold change between severe PE and controls; the y-axis is negative log-transformed p-values after BH adjustment. The red dots are differentially methylated probes (DMP) associated with severe PE after BH adjustment, whereas the black dots represent non-significant probes. (C) The volcano plot after adjusting for all confounding factors.

As a comparison, we first conducted differential methylation analysis on severe PE without adjusting for any clinical confounders or cell-type proportions. The analysis reveals a global hypomethylation pattern (**Fig. 4B**). We identified 229,730 differentially methylated CpGs with adjusted p-values less than 0.05. Among these CpGs, 184,102 exhibited hypomethylation, while 45,628 displayed hypermethylation. However, when we redid the differential methylation analysis after adjusting for cell type heterogeneity and patient characteristics, all the CpGs differentially methylated above are no longer significant, except for a single CpG site labeled as cg20135196 on Chromosome 6, with FDR=0.014 (**Fig. 4C**). Interestingly, this locus is rarely reported before. A total of 11 genes (**Supple Table 8**) are identified as the nearest genes to this locus on both sides. ZNF 184 is the closest gene by distance. It is a zinc finger protein involved in transcription regulation. A series of genes of nucleosome components are located on the opposite side of ZNF 184.

Additionally, we extended the differential methylation analysis to the gene level. We aggregated the CpGs located on gene promoters as the representation of promoter-level methylation (see Methods). Before adjusting for confounders, we detected 4,767 differentially methylated genes. However, upon adjusting for both clinical variables and cell types, none of the genes exhibited statistical significance. Similarly, we conducted a differential methylation analysis at the pathway level, employing the Pathifier algorithm (see **Methods)**. Before the confounding adjustment, we detected 200 significant pathways. Nevertheless, after accounting for confounders, none of the pathways remained significant. In conclusion, the most concerning finding is that the observed DNA methylation variation among the whole cord blood samples is primarily associated with cell type heterogeneity rather than severe PE.

To confirm the significant influence of confounders on EWAS associated with severe PE, we re-analyzed the Illumina 450k DNA methylation data from Ching et al, which were obtained from different samples^16^. Using the original analysis pipeline that did not consider clinical confounding, we reproduced the differential methylation results earlier, which reported 68,458 significant CpGs (**Supple Fig. 3A**). Subsequently, we estimated the cell type proportions using Houseman’s CP algorithm and the new combined cord blood reference reported in this study. We then conducted the SOV analysis by considering cell proportions and clinical variables. We identified cell type proportions, gestational age, maternal age and PE as significant confounders, as they have F-statistics > 1 (**Supple Fig. 3B**). Consistent with the observation on the EPIC array cord blood dataset earlier, the dataset by Ching et al. yields no significant CpG (**Supple Fig. 3C**) once adjusted for these confounders. In summary, using two cord blood datasets, we demonstrated that confounding effects from clinical variables and cell type heterogeneity are common challenges that need to be addressed by EWAS analysis in cord blood.

### Association between cord blood cell type and gestational age

Our earlier analysis shows that estimated cell proportions in cord blood are correlated with gestational age (**Fig. 3C**). We thus conducted a more in-depth analysis. The most noticeable correlation comes from granulocytes, whose proportions increase from around 25% in week 32 to over 50% in week 40, with P=0.00015 (**Fig. 5A**). The proportions of monocytes also significantly increase as the gestation progresses, after adjusting for other variables (p=0.0051, **Fig. 5B**). On the contrary, nRBC, natural killer, and B cell significantly decrease along the gestation (p = 6.62e-6; p=0.0054; p=0.0087, **Fig. 5C-E**). These trends of changes are maintained when the samples are stratified into case and control groups (**Supple Fig. 4**), without significant interaction between each cell proportion and case/control labels when regressing them on gestational ages (y-values).

**Fig. 5:**
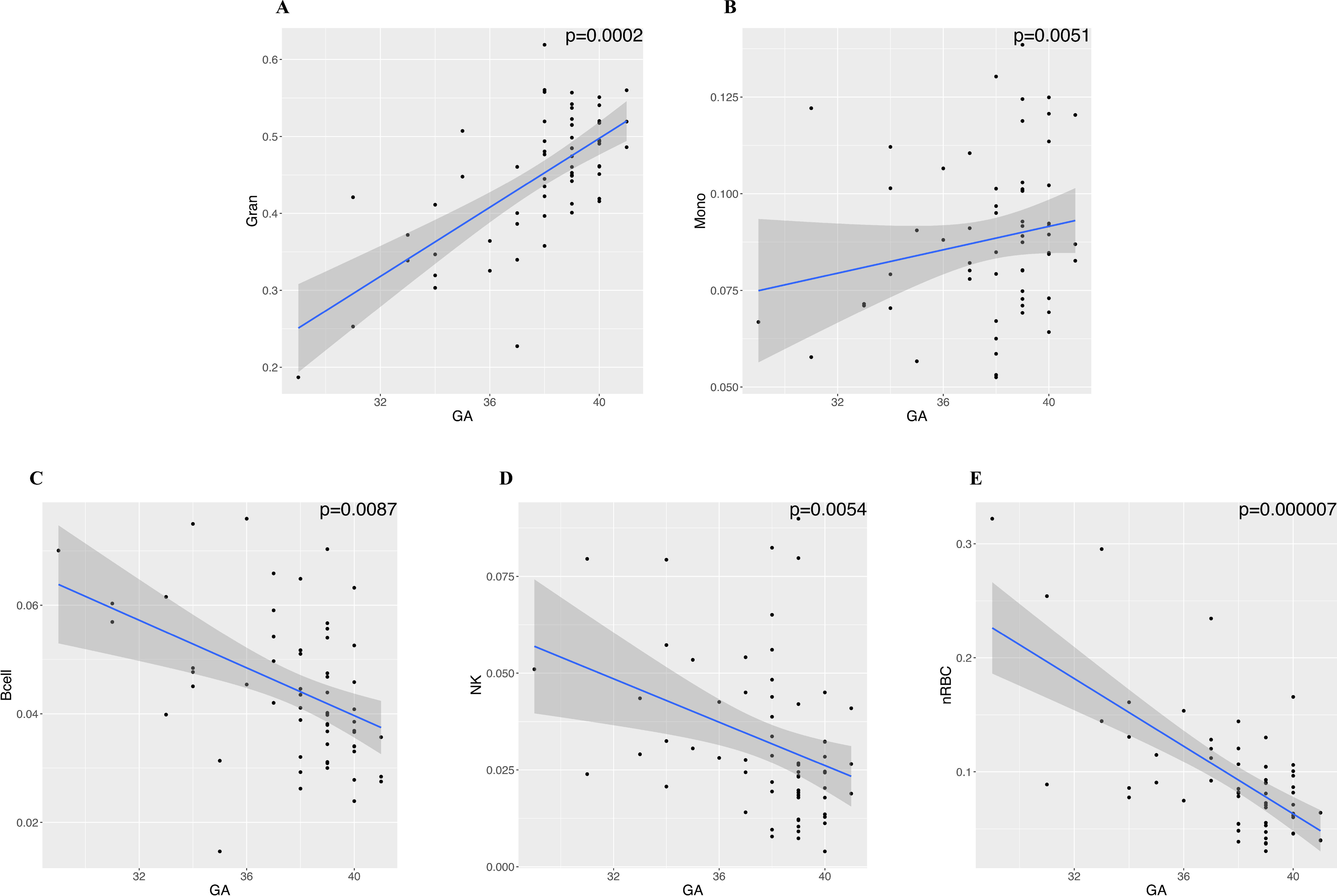
Cell type proportion changes with gestational age. Scatter plots labeled (A) to (E) depict the changes in the proportions of each cell type in cord blood along with gestational age. The reported p-value measures the relationship between GA and each cell type, with a threshold of p-value < 0.05.

Furthermore, we validated the trends of cell type proportion through gestational age using another public cord blood peripheral blood mononuclear cell (PBMC) Illumina HumanMethylation450 BeadChip methylation dataset (GEO accession ID: <u>GSE110828</u>), which comprises 20 PE cases and 90 non-PE controls^41^. Both the case and control groups include large percentages of preterm samples, with the delivery gestational ages ranging from 26.14 to 38.14 in cases and 23.00 to 41.29 in controls. We deconvoluted the PBMC cell types with Houseman’s CP method using the new reference data we created here without the granulocytes due to their absence in PBMC. We performed linear regression of gestational age over the cell type, severe PE and their interaction terms similar to those in **Fig. 5** We observe the same increasing trend for monocytes, and the same decreasing trend for nRBC cell, natural killer cell and B cell (**Supple Fig. 5**). Again, none of the interaction terms between severe PE and gestational age turned out to be significant, suggesting that their effects on cell proportion changes are independent.

## Discussion

In this study we showed and validated the importance of confounding adjustment in bulk DNA methylation analysis on the EWAS result, using two datasets on cord blood samples exposed to severe PE. As the most significant confounder among clinical variables, gestational age affects various blood cell type proportions (eg. granulocyte, nRBC, CD4T and B cells). We showed that many CpGs, genes and pathways will be artificially and wrongfully detected if we do not adjust for the profound confounding due to cell type composition and other clinical variables such as gestational age. Despite the lack of CpG changes to severe PE, many cell types’ proportion changes do differ between severe PE vs. controls. Thus, we conclude the major effect of PE on cord blood is not through CpG methylation changes, but the proportion changes in different cell types.

Among all confounders in EWAS analysis, cell type heterogeneity is one of the most common and important confounders faced by researchers. Over the years, various cell type estimation methods for bulk-level epigenetic data have been developed ^35,40^, which made the assessment of cell type confounding effects possible. In 2013, Liu et al. first reported a large reduction in differentially methylated probes related to Rheumatoid Arthritis after adjusting for cell type composition in whole blood^42^. Some later studies confirmed the effect of cell type heterogeneity on EWAS research in other tissues such as breast tissue, saliva, and placenta tissue^8,43,44^. Unlike many other diseases, pregnancy complications in the form of severe PE are unique in terms of confounding. Many fetuses have to be delivered preterm, leading to the case group with smaller GA compared to the healthy full-term controls. Therefore, gestational age and PE are two conditions which go hand-in-hand, and gestational age is a major confounder for studying severe PE. More importantly, the cell proportions in severe PE case groups are also affected by the disease itself, making them critical confounders in EWAS studies. Although several previous studies aimed to identify PE-related epigenetic biomarkers using cord blood samples^17–20^, the importance of adjusting for cell type heterogeneity was mostly (3 out of 4) overlooked. This could have led to biased results or even false biomarkers. This could also be one of the reasons for the inconsistency in these previous studies.

Furthermore, we noticed consistent associations between cell type compositions in cord blood and gestational age, both in severe PE cases and controls. Granulocyte proportion showed the strongest positive association with gestational age, agreeing with the previous findings that granulocyte in the fetus increases drastically in the last trimester of pregnancy^41^. The estimated proportion of nRBC in our study decreases as gestational age increases, also consistent with previous findings^37–39^. Additionally, elevated nRBC was also found in preterm infants and infants with lower birth weight^45^, providing additional supporting evidence to our finding. We also observed a significant increase in monocytes and significant decreases in B cells and NK cells as gestational age increases. Taken together, these findings probe into the dynamic nature of cell type composition in cord blood during gestation. Future EWAS studies utilizing cord blood samples collected at different gestational ages should carefully adjust for cell type heterogeneity.

## Conclusion

In summary, we found the lack of evidence for significant CpG methylation changes in EWAS analysis in association with severe PE, after adjusting for cell type heterogeneity and clinical variables such as gestational age. Wrongful and artificial CpG methylation biomarkers would have been obtained without such adjustment. Instead of altering CpG methylations, severe PE is associated with significant changes in several cell proportions in the cord blood. We also showed that many cell types’ proportions change drastically during pregnancy.

## Data Availability

All data produced in the present study are available upon reasonable request to the authors

## Author Contributions

LXG conceived this project and supervised the study. WL and XY contributed equally to data analysis, result generation, and manuscript writing. ZM and YD assisted in the data processing. CL, FMA, and PAB contributed to sample collection, coordination and the experimental design of the DNA methylation. All authors have read, revised and approved the manuscript.

## Acknowledgment

We thank the Genomics Shared Resources of the University of Hawaii Cancer Center for performing the methylation assays.

## Materials & Correspondence

Correspondence to Lana X Garmire

## Supplementary Figures

**Supplementary Figure 1: Data Processing Workflow.** The complete data pre-processing procedures consisted of filtration, imputation of missing values, quality control checks, normalization, batch correction, singular value decomposition analysis, and conversion of beta values to M-values.

**Supplementary Figure 2: Data Quality Control.** (A) and (B) Density plots for before and after the removal of one control sample with a distinct beta density distribution. (C) and (D) Plots of the singular value decomposition analysis are presented before and after the removal of batch effects.

**Supplementary Figure 3: Validation of the impact of confounding adjustment using Ching et al.’s 450k cord blood methylation data**^17^. (A) The volcano plot of differential methylation results without confounder adjustment as done by Ching et al. using their data. The red dots are differentially methylated probes (DMP) associated with EOPE after BH adjustment, whereas the black dots represent non-significant probes. (B) The Source of Variance (SOV) analysis was conducted on both clinical variables and cell types. Confounding factors were identified by considering variables with an F-mean value greater than 1. (C) The volcano plot after adjusting for all confounding factors.

**Supplementary Figure 4: Cell type changes with gestational age by sample group.** Scatter plots labeled (A) to (G) depict the changes in the proportions of each cell type along with gestational age. The green line in each plot represents the PE case group, while the red line represents the control group. The reported p-value measures the interaction between GA and the sample group of trends between the PE case group and the control group, with a threshold of p-value < 0.05. A non-significant p-value indicates the trends between cell proportion and GA are consistent in the case and control groups.

**Supplementary Figure 5: Cell proportion changes with gestational age are consistent in different datasets.** The scatter plots compare the changes in cell-type proportions with gestational age in two datasets. Plots (A - F) display the comparisons within PE case samples for both studies, while plots (G) through (L) display the comparisons within control samples for both studies. The purple line in each plot represents the cell proportion in our whole cord blood samples, while the orange line represents the cell proportion in PBMC cord blood samples from another study. The p-values of the interaction term between GA and datasets are reported in each plot. A non-significant p-value suggests the trend between cell proportion and GA is consistent in the two datasets.

## Supplementary Tables

**Supplementary Table 1: Linear regression of each cell type on clinical variables**

**Supplementary Table 2: The closest functional genes to the significant CpG site, cg20135196**

