## Supplementary Materials for "Severe preeclampsia is not associated with significant DNA methylation changes but cell proportion changes in the cord blood - caution on the importance of confounding adjustment"

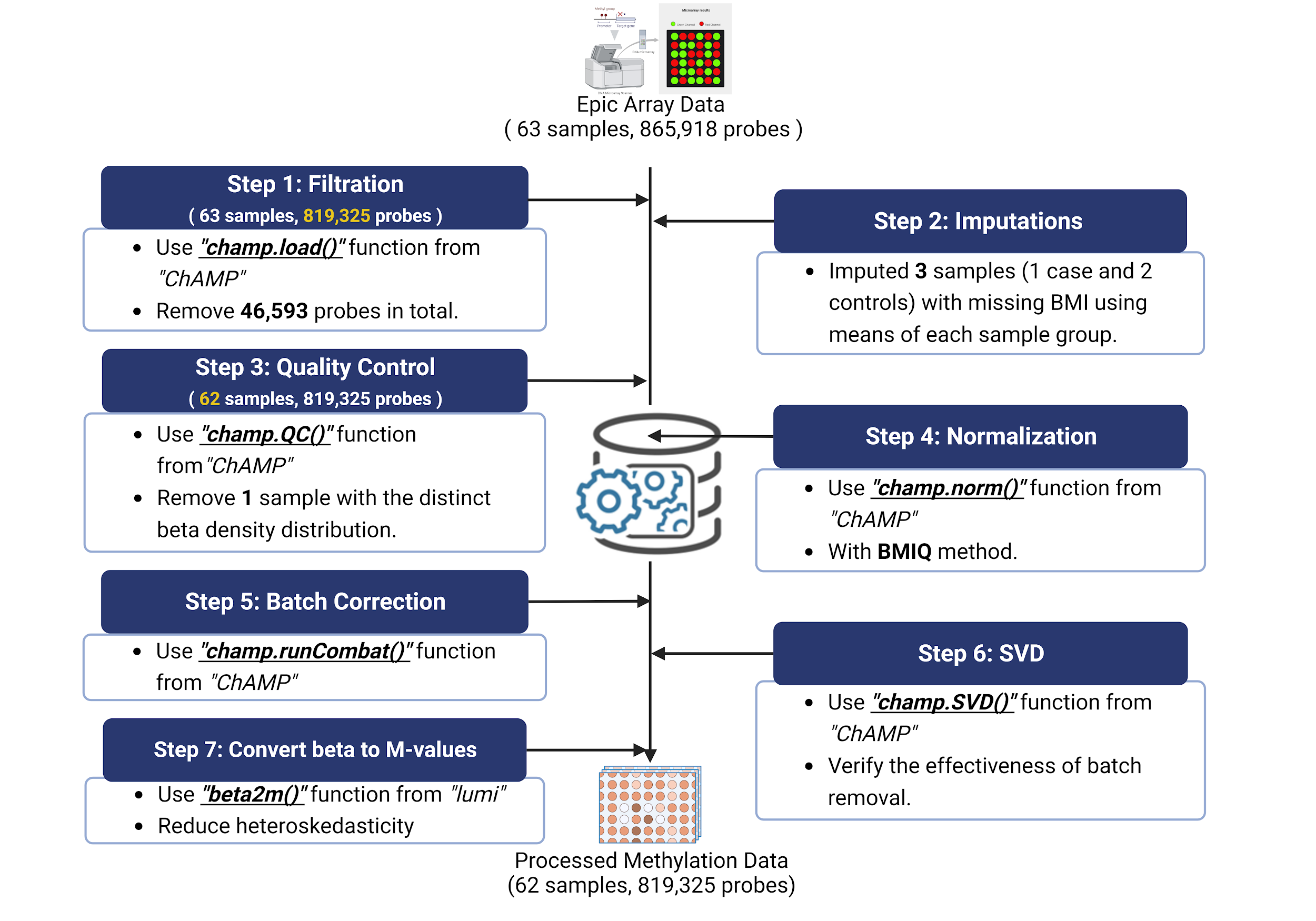

**Supplementary Figure 1:** Data Processing Workflow. The complete data pre-processing procedures consisted of filtration, imputation of missing values, quality control checks, normalization, batch correction, singular value decomposition analysis, and conversion of beta values to M-values.

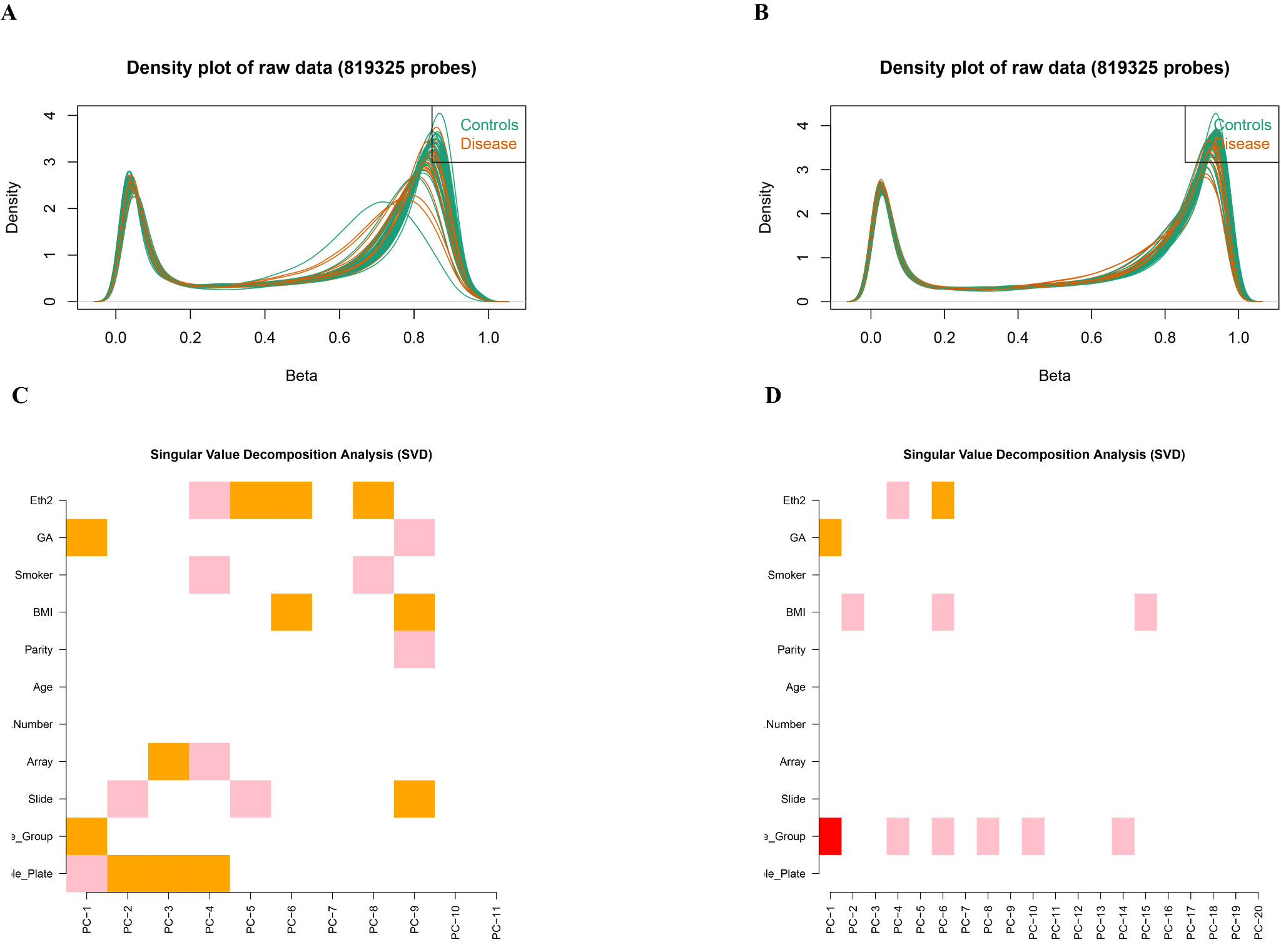

**Supplementary Figure 2: Data Quality Control.** (A) and (B) Density plots for before and after the removal of one control sample with a distinct beta density distribution. (C) and (D) Plots of the singular value decomposition analysis are presented before and after the removal of batch effects.

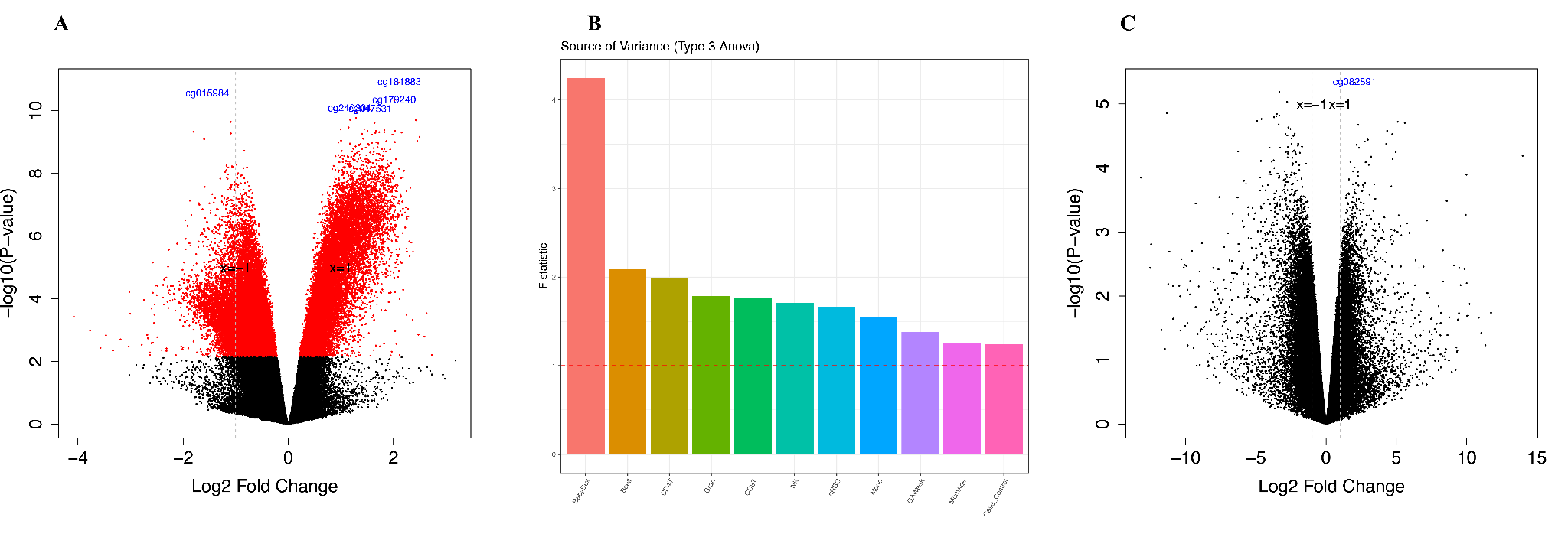

**Supplementary Figure 3: Validation of the impact of confounding adjustment using Ching et al.’s 450k cord blood methylation data.** (A) The volcano plot of differential methylation results without confounder adjustment as done by Ching et al. using their data. The red dots are differentially methylated probes (DMP) associated with EOPE after BH adjustment, whereas the black dots represent non-significant probes. (B) The Source of Variance (SOV) analysis was conducted on both clinical variables and cell types. Confounding factors were identified by considering variables with an F-mean value greater than 1. (C) The volcano plot after adjusting for all confounding factors.

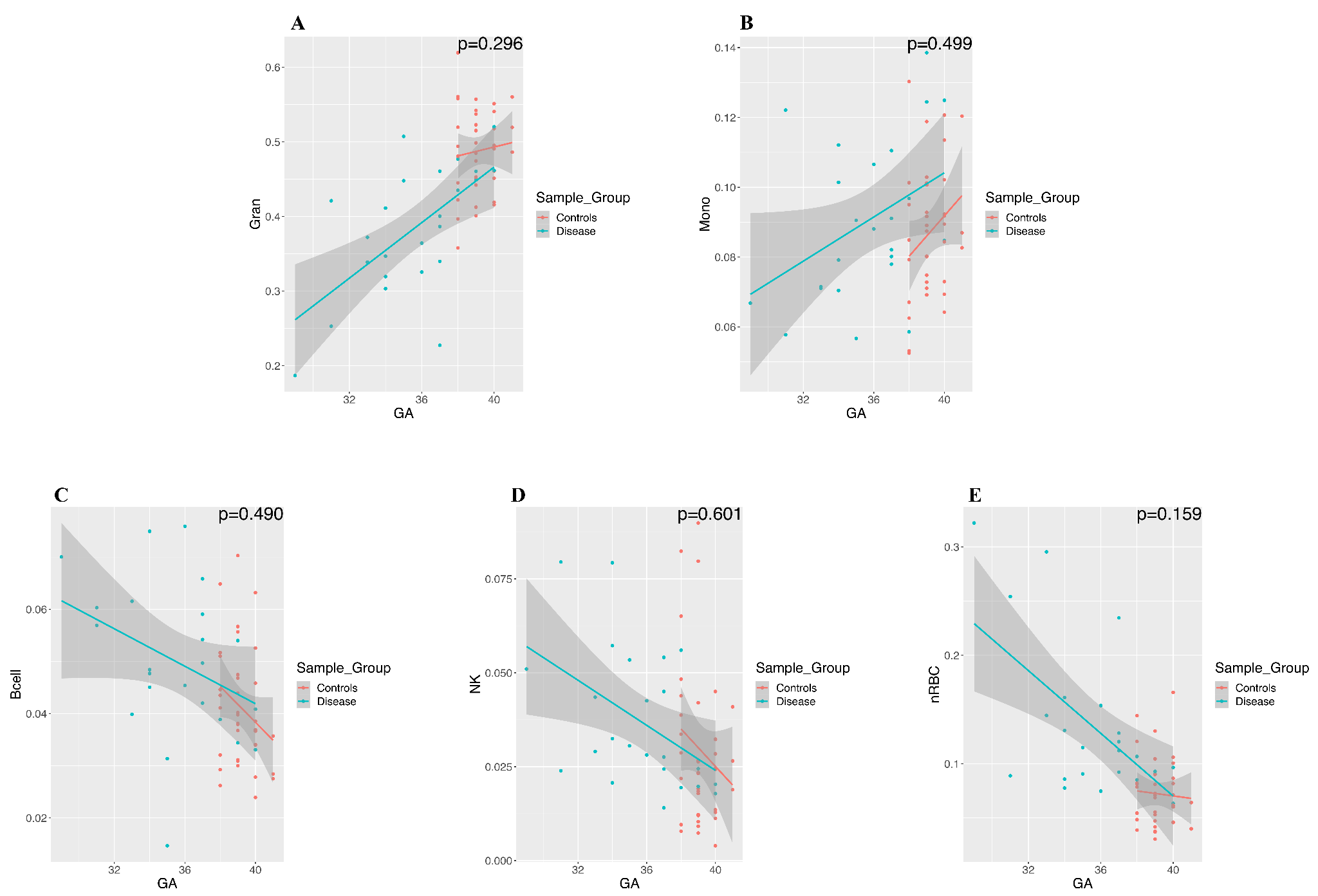

**Supplementary Figure 4: Cell type changes with gestational age by sample group.** Scatter plots labeled (A) to (G) depict the changes in the proportions of each cell type along with gestational age. The green line in each plot represents the PE case group, while the red line represents the control group. The reported p-value measures the interaction between GA and the sample group of trends between the PE case group and the control group, with a threshold of p-value < 0.05. A non-significant p-value indicates the trends between cell proportion and GA are consistent in the case and control groups.

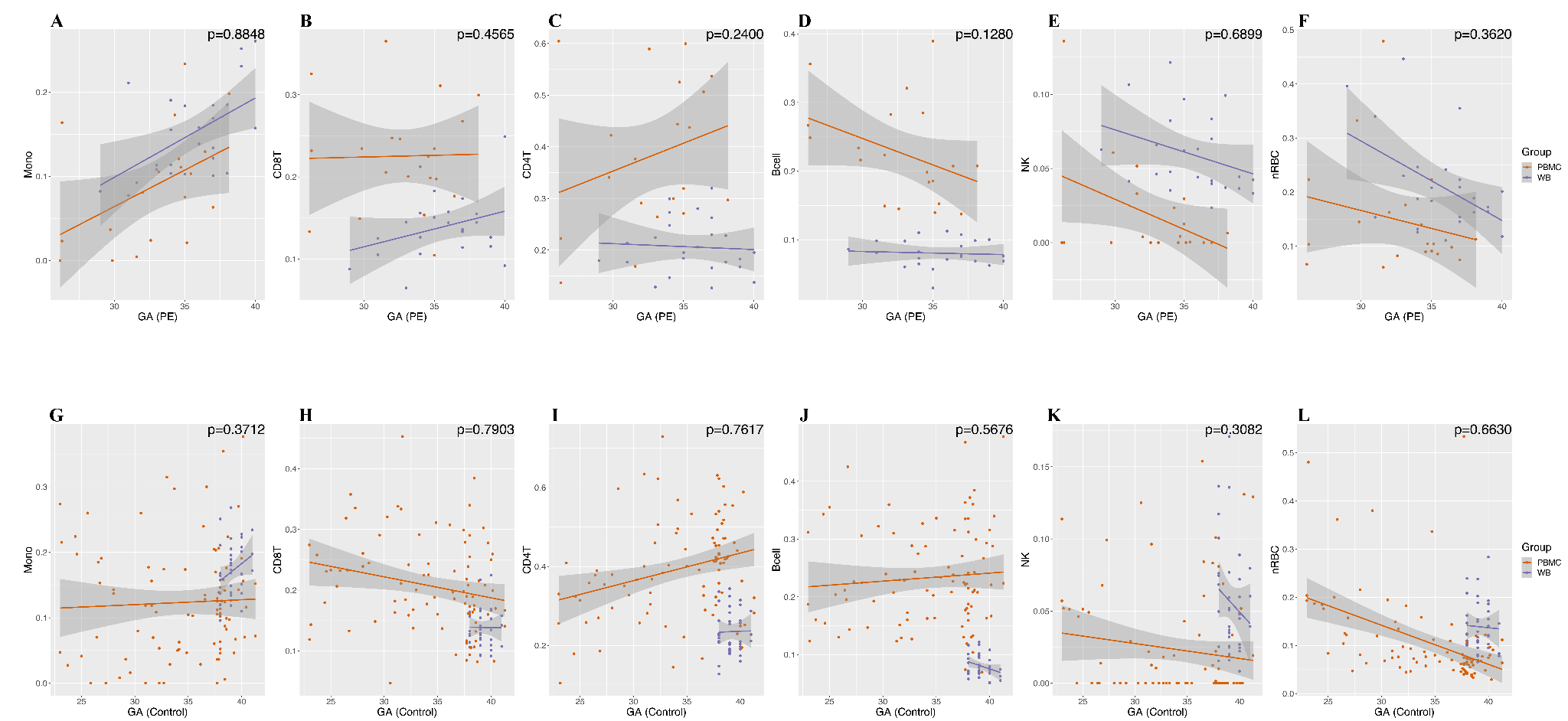

**Supplementary Figure 5: Cell proportion changes with gestational age are consistent in different datasets.** The scatter plots compare the changes in cell-type proportions with gestational age in two datasets. Plots (A - F) display the comparisons within PE case samples for both studies, while plots (G) through (L) display the comparisons within control samples for both studies. The purple line in each plot represents the cell proportion in our whole cord blood samples, while the orange line represents the cell proportion in PBMC cord blood samples from another study. The p-values of the interaction term between GA and datasets are reported in each plot. A non-significant p-value suggests the trend between cell proportion and GA is consistent in the two datasets.

**Supplementary Tables**

**Supplementary Table 1: Linear regression of each cell type on clinical variables**

| **Supplementary Table 1: Linear regression of each cell type on clinical variables** | | | | | | | | | | | | | | |
| --- | --- | --- | --- | --- | --- | --- | --- | --- | --- | --- | --- | --- | --- | --- |
|  | *CD8T* | | *CD4T* | | *B cell* | | *Granulocyte* | | *Monocyte* | | *Natural Killer* | | *nRBC* | |
|  | *Coefficient* | *P-value* | *Coefficient* | *P-value* | *Coefficient* | *P-value* | *Coefficient* | *P-value* | *Coefficient* | *P-value* | *Coefficient* | *P-value* | *Coefficient* | *P-value* |
| ***(Intercept)*** | *0.063* | *0.302* | *0.229* | *0.068* | *0.148* | ***3.85E-04*** | *-0.278* | *0.162* | *-0.097* | *0.125* | *0.207* | ***0.001*** | *0.664* | ***3.71E-06*** |
| ***PE*** | *0.02* | ***0.007*** | *0.002* | *0.917* | *0.001* | *0.844* | *-0.042* | *0.071* | *0.016* | ***0.029*** | *-0.005* | *0.519* | *0.006* | *0.689* |
| ***GA*** | *0.001* | *0.665* | *-0.003* | *0.378* | *-0.002* | ***0.009*** | *0.019* | ***0*** | *0.004* | ***0.005*** | *-0.004* | ***0.005*** | *-0.015* | ***6.62E-06*** |
| ***BMI*** | *-0.001* | ***0.002*** | *-0.001* | *0.131* | *-1.76E-04* | *0.42* | *0* | *0.81* | *3.62E-04* | *0.299* | *0* | *0.864* | *0.002* | ***0.007*** |
| ***Caucasian*** | *0.012* | *0.086* | *0.002* | *0.907* | *0.007* | *0.116* | *0.001* | *0.975* | *-0.011* | *0.143* | *0.001* | *0.896* | *-0.013* | *0.406* |
| ***Pacific Islander*** | *0.018* | *0.013* | *0.014* | *0.338* | *0.006* | *0.191* | *0.025* | *0.277* | *-0.009* | *0.237* | *-0.011* | *0.139* | *-0.044* | ***0.005*** |
| ***Parity*** | *0.002* | *0.375* | *0.004* | *0.228* | *0.001* | *0.245* | *-0.004* | *0.51* | *0.001* | *0.542* | *-0.001* | *0.5* | *-0.003* | *0.483* |
| ***Age*** | *0* | *0.764* | *0* | *0.746* | *-3.80E-04* | *0.256* | *0.002* | *0.292* | *4.14E-04* | *0.437* | *-0.001* | *0.229* | *-0.002* | *0.133* |

**Supplementary Table 2: The closest functional genes to the significant CpG site, cg20135196**

| **Gene position** | **Gene name** | **Gene function** |
| --- | --- | --- |
| 138,254 bp away from left side | ZNF 184 | •May be involved in transcriptional regulation |
| 699,912 bp away from right side | H2BC13 / H2AC13 / H2AC14 / H2BC14 / H4C11 / H4C12 / H2AC15 / H2BC15 / H2AC16 | •Core component of nucleosome.  •Nucleosomes wrap and compact DNA into chromatin, limiting DNA accessibility to the cellular machineries which require DNA as a template.  •Histones thereby play a central role in transcription regulation, DNA repair, DNA replication and chromosomal stability.  •DNA accessibility is regulated via a complex set of post-translational modifications of histones, also called histone code, and nucleosome remodeling. |
|  | H1-5 | •Histone H1 protein binds to linker DNA between nucleosomes forming the macromolecular structure known as the chromatin fiber.  •Histones H1 are necessary for the condensation of nucleosome chains into higher-order structured fibers.  •Acts also as a regulator of individual gene transcription through chromatin remodeling, nucleosome spacing and DNA methylation (By similarity). |

** Gene position and Gene name are from “UCSC Genome Browser on Human (GRCh37/hg19)”**^45^**;*

** Gene function is from “GeneCards”**^46^**.*
